# Sentiment in Clinical Notes: A Predictor for Length of Stay?

**DOI:** 10.64898/2026.03.16.26348553

**Authors:** Aidan Boyne, Maximillian Feygin, John Sholeen, Andrew Zimolzak

## Abstract

**Background:** Length of stay (LOS) is a critical metric for hospital operational efficiency. While structured clinical data is widely used to predict LOS, unstructured admission notes contain latent prognostic information regarding diagnostic uncertainty and disease complexity. This study evaluates the efficacy of extracting sentiment and direct LOS estimates from admission notes to predict patient hospitalization duration.

**Methods:** We conducted a retrospective study of 4,503 adult patients admitted with community-acquired pneumonia between 2013 and 2023. Admission history and physical notes were preprocessed and filtered to extract physician-generated narratives. We evaluated four natural language processing models, VADER, TextBlob, Longformer, and an open-source large language model (GPT-oss-20B), to generate zero-shot sentiment scores. Additionally, GPT-oss-20B was prompted to directly estimate LOS. Model outputs were correlated with actual LOS using linear regression and Pearson correlation coefficients.

**Results:** Sentiment models demonstrated statistically significant, albeit weak, correlations with actual LOS. Longformer achieved the highest variance explained among sentiment classifiers (R^2^ = 0.019). Direct LOS estimation by the LLM outperformed sentiment-based approaches, demonstrating the strongest correlation with actual hospital duration (r = -0.218, p < 0.001). Model agreement was generally poor (ICC = 0.059), and computational time varied drastically, from 2.6 seconds per 100 notes (TextBlob) to over 370 seconds (GPT-oss-20B).

**Conclusion:** Zero-shot sentiment analysis of clinical notes yields a small but measurable correlation with LOS, limited primarily by the objective, non-evaluative nature of clinical documentation. Direct LLM estimation of clinical outcomes outperforms emotional sentiment extraction. Future predictive systems should integrate computationally efficient NLP models capable of capturing latent clinical complexity alongside established structured data variables.

## 1. Introduction

Length of stay (LOS) is a core metric used by medical institutions to evaluate operational efficiency and quality of care. Predicting LOS at the time of admission remains difficult, and existing models rely heavily on structured variables such as patient demographics, admission vitals and labs, and comorbidity indices^1^. Unstructured data, particularly the clinical note, is a potential source of additional prognostic information reflecting physician judgement, diagnostic uncertainty, and disease complexity. Recent work has demonstrated that unstructured documentation often contains underlying signals independent from structured data that can predict patient outcomes^2^. Sentiment analysis, or the process of computationally identifying negative, neutral, or positive attitude in text, is a particularly promising method to extract these signals.

Initially gaining traction in the finance sector, where it was used to analyze public sentiment toward publicly traded companies on Twitter and other chat forums to inform trading strategy, sentiment analysis has since been applied in a variety of contexts, including medicine. Prior sentiment analysis studies primarily used rules-based tools to assess mortality risk in critical care populations and were largely focused on a single de-identified database of patient notes, MIMIC-III. Weissman et al. demonstrated variable construct validity across six rule-based sentiment tools in ICU notes^3^. Others have applied sentiment scoring to lung cancer patients in the Veterans Affairs system and evaluated nursing note sentiment to predict 28-day mortality in sepsis^4,5^. These studies indicate that sentiment is detectable and sometimes predictive, but they are constrained by single disease contexts, single data sources, and reliance on older rule-based approaches.

More recently, large language models (LLMs) have shown promise as effective sentiment classifiers both for general text and demonstrated high performance in medical natural language processing (NLP) tasks, including outcome prediction^6,7^. However, it has been shown that LLMs are not necessarily superior to older NLP techniques such as the rules-based VADER and TextBlob models or encoder models such as Longformer^8^.

In this study, we evaluate whether sentiment derived from admission using a variety of models can predict LOS in adult patients admitted with community-acquired pneumonia (CAP). We compare rule-based, encoder-based, and LLM-based sentiment classifications, analyze their relationship to LOS and potential confounders, and test whether sentiment offers predictive value. To our knowledge, we are the first to examine LLM-driven sentiment extraction from clinical notes as a predictor of LOS and to compare LLM sentiment extraction in a clinical context head-to-head with other NLP techniques.

## 2. Methods

### 2.1 Dataset Preparation

Extraction of the data used in the study was determined to be exempt from IRB review. Admission history and physical (H&P) notes were extracted from the Baylor St. Luke’s Medical Center electronic health record system (Epic) for all patients aged ≥18 years admitted with a diagnosis of CAP between June 2013 and June 2023. For each encounter, admit and discharge timestamps, demographic information, and author credentials were collected. Only notes written by staff physicians, fellows, or residents were included. The full text of each of these notes and associated information comprised the first dataset, referred to as the full-text dataset for the remainder of this work.

### 2.2 Preprocessing

Text was standardized through lowercasing and whitespace normalization. A subset of notes underwent manual review to identify phrases most likely to contain physician-generated narrative text. These phrases were then used to filter relevant information (e.g., history and physical, assessment, plan) from auto-generated or filler text using fuzzy regex matching. Before being fed to each model, filtered text exceeding the maximum input length was divided into chunks, taking care to preserve sentences if possible.

### 2.3 Sentiment Classification

Sentiment scores ranging from -1 (most negative) to 1 (most positive) were generated using several approaches, all implemented in Python version 3.12. The rules-based VADER^9^ and TextBlob models and the encoder-based Longformer^10^ model were used to generate scores for both the full-text and filtered-text databases (**Figure 1**). GPT-oss-20B^11^, an open source LLM created by OpenAI, was run on our local machine to preserve patient privacy using LM Studio and used to classify only the filtered-text database using the prompt:

> *“I am conducting a project to analyze the sentiment of medical notes. You are an impartial and objective rater. Rate the sentiment of this text from -1 (negative, unfavorable for patient) to 1 (positive, favorable for patient). Reply with only the number. The first output you provide should be a number between -1 and 1 inclusive, and you must limit your output to a maximum of 5 characters (e*.*g*., *if you wanted to output 0*.*184321, you would stop at 0*.*184)*.*”*
>
> *“Here is the clinical text you are rating: {NOTE TEXT HERE}” “Rating:”*

**Figure 1.**
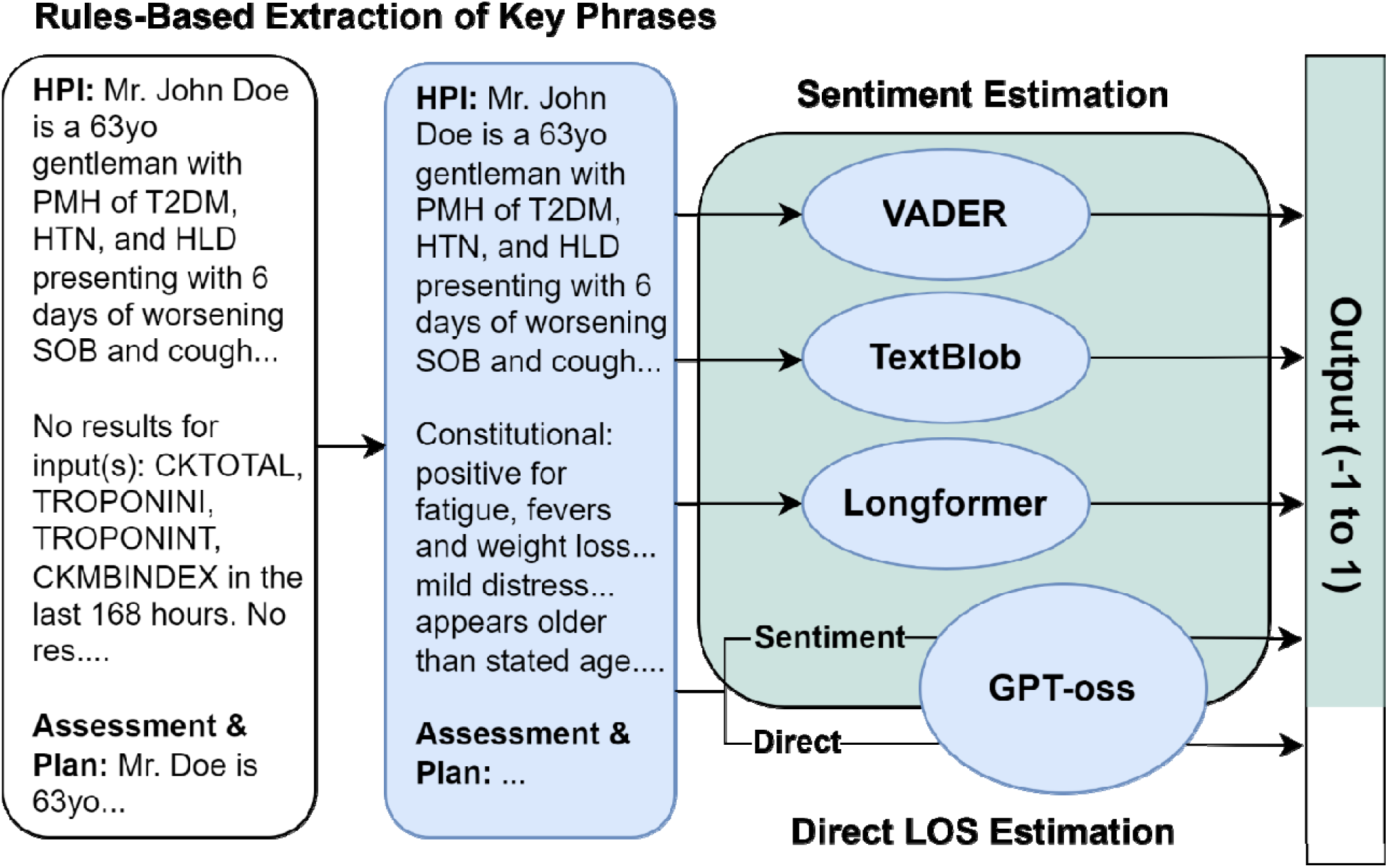
Sentiment and Direct Length of Stay (LOS) Scoring. Each full note is passed to VADER, TextBlob, and Longformer models pre-trained for sentiment analysis. The full notes are then passed through a filter that extracts key phrases of the note, such as the history and physical, assessment, and plan. This step reduces noise such as text from repeated note template phrases or unfilled values. The filtered notes are passed to the VADER, TextBlob, and Longformer, and GPT-oss for sentiment estimation and again to GPT-oss for direct LOS estimation. Text is split into chunks at sentence boundaries if it exceeds the maximum input length of the model. Outputs are normalized to a scale of - 1 (most negative sentiment) to 1 (most positive sentiment).

When note text exceeded a model’s maximum input length, the sentiment scores from each chunk were averaged, ignoring any scores of 0. Model inference was performed on a local workstation equipped with 16 GB of system RAM and an NVIDIA RTX A6000 GPU with 48 GB of VRAM for hardware acceleration.

### 2.4 Zero-shot LOS Estimation

To evaluate the effect of prompt language on the predictive value of model output, GPT-oss was also used to estimate LOS directly from note text using the prompt:

> *“I am conducting a project to predict the length of stay (LOS) of patients admitted for pneumonia based on their admission note. You are an impartial and objective medical administrator tasked with predicting the LOS. Return the expected LOS for this admission from -1 (very long expected LOS) to 1 (very short expected LOS). Reply with only the number. The first output you provide should be a number between -1 and 1 inclusive and you must limit your output to a maximum of 5 characters (e*.*g. if you wanted to output 0*.*184321, you would stop at 0*.*184)*.*”*
>
> *“Here is the admission note for your rating: {NOTE TEXT HERE}” “Rating:”*

### 2.5 Outcomes

LOS was calculated as discharge time minus admission time. Linear regressions for each model output and LOS were conducted, and the degree of explainable variability was reported as R^2^. Pearson correlation was calculated between model outputs (sentiment scores or directly estimated LOS) and the actual LOS, adjusting for multiple comparisons using the Benjamini-Hochberg method and a significance threshold of α = 0.05. Model agreement was calculated using the interclass correlation coefficient (ICC) and interpreted according to the criteria established by Koo and Li^12^.

## 3. Results

### 3.1 Dataset

A total of 4530 unique admission notes authored by medical doctors for patients hospitalized for a primary diagnosis of CAP between June 2013 and June 2023 were extracted. After filtering LOS outliers, 4503 patients were included in the analysis. Of the 4503 patients, 2492 (55.3%) were male and 2011 (44.7%) were female. The mean and standard deviation of patient age were 64.0 ± 18.0 years, and the median length of stay was 9.0 days (interquartile range 5.0, 19.0 days).

### 3.2 Correlations with LOS

Among the four sentiment predictors, Longformer achieved the strongest fit with linear regression (R^2^ = 0.019), followed by Vader (R^2^= 0.014), and the LLM (R^2^= 0.008). TextBlob did not explain any variability in LOS as determined by linear regression (R^2^ = 0.000) (**Figure 2**). Direct LOS estimation by the LLM reached an R^2^ value of 0.017. Correlation as measured by the Pearson coefficient followed a similar pattern: direct LLM estimation was most correlated (r = -0.218, p < 0.001), followed by Vader (r = 0.170, p <0.001), Longformer (r = -0.119, p < 0.001), LLM sentiment (r = - 0.118, p<0.001), and TextBlob (r = -0.030, p = 0.041) (**Table 1**).

**Table 1.** Results of sentiment and direct length of stay (LOS) estimation. Correlation of model estimates with actual LOS is reported using the Pearson correlation coefficient with Benjamini-Hochberg correction for multiple comparisons.

| Model | Mean Estimate<br>(±SD) | LOS<br>Correlation | p (corrected) | Time per 100<br>notes |
| --- | --- | --- | --- | --- |
| Vader | -0.364 ± 0.428 | 0.170 | < 0.001 | 3.1s |
| TextBlob | 0.016 ± 0.103 | -0.030 | 0.041 | 2.6s |
| Longformer | -0.396 ± 0.372 | -0.119 | < 0.001 | 9.0s |
| LLM<br>(Sentiment) | -0.165 ± 0.278 | -0.118 | < 0.001 | 374.0s |
| LLM (Direct) | -0.325 ± 0.480 | -0.218 | < 0.001 | 344.7s |

**Figure 2.**
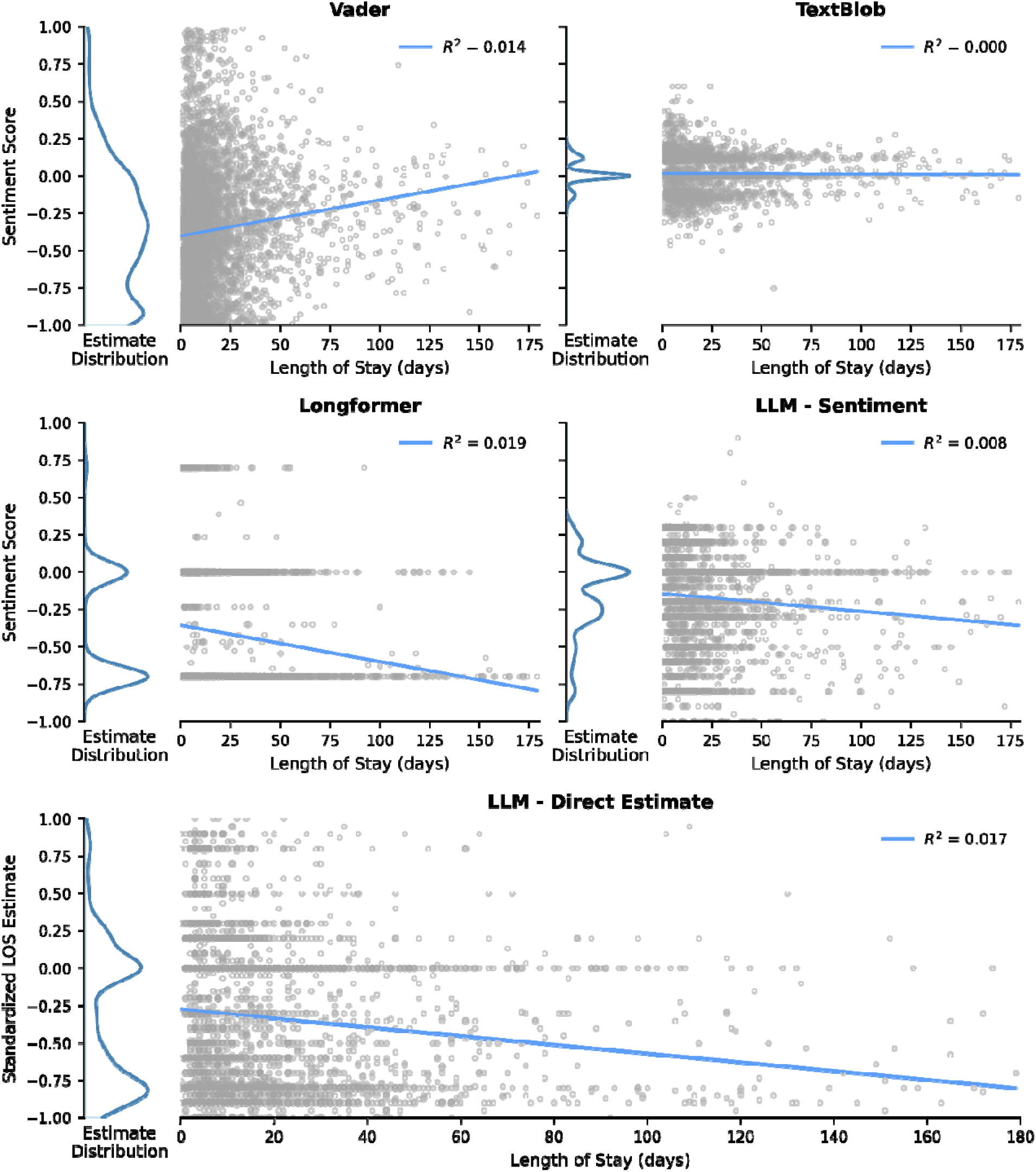
Linear regression of model outputs on length of stay (LOS). Distributions of model estimates are plotted to the left of each regression. The line of best fit for each regression is superimposed on the corresponding scatterplots of the estimate/LOS pairs. The coefficient of determination for the regressions is provided in the upper right corner of each plot.

### 3.3 Sentiment Distribution, ICC, and Computation Time

On average, four of the five models estimated a moderately low sentiment (sentiment models) or a long length of stay (direct estimator), while TextBlob estimated a neutral sentiment. Distribution of sentiment and direct LOS estimation by the models given in **Table 1**. Model agreement evaluated by a two-way random effects model was low. The single-measure agreement ICC(2,1) was 0.059 [95% CI: 0.061, 0.083], while average agreement across all five models ICC(2,k) was 0.280, indicating a poor level of agreement between models. The time required to process 100 filtered clinical notes varied from 2.6 seconds for the TextBlob model to over 370 seconds for GPT-oss 20B (**Table 1**).

## 4. Discussion

In this study, we evaluated the utility of sentiment analysis and LLM-based estimation for predicting LOS based on unstructured clinical text. While sentiment scores as estimated by most models showed a statistically significant correlation with LOS, the magnitude of this correlation was notably small. From a practical perspective, these findings suggest that while a relationship between admission note sentiment and LOS exists, the predictive utility of sentiment alone as a biomarker for hospital stay is likely limited. Admittedly, estimation of LOS based off a patient’s state at admission is a challenging problem, as patient trajectories are influenced by myriad factors that unfold dynamically during hospitalization.

### 4.1: The Signal-to-Noise Challenge

The modest performance of sentiment models demonstrated in this study likely stems from a variety of sources. First, clinical documentation is often filled with autogenerated phrases, imported laboratory values, and copy forwarded text that contain no useful prognostic information or measurable sentiment^13^. Though we attempted to address this through rules-based filtering and fuzzy regex matching, some irrelevant information inevitably bypasses these filters and dilutes any sentiment information.

Even in the absence of irrelevant “note bloat”, the objective nature of clinical documentation often results in text with very little detectable sentiment. Though writing styles vary, physicians are trained to write descriptive, objective narratives. Words that are clinically severe (e.g., “septic”, “intubated”, “hypotensive”) do not necessarily carry the same negative linguistic valence in standard pre-trained models as words indicating frustration or anger. Consequently, traditional sentiment measuring positive or negative emotionality is often a poor proxy for clinical severity and prognostic risk.

### 4.2 Latent Information in Unstructured Text

Despite the challenges with little sentiment information and large amounts of noise in clinical notes, the performance of certain models suggests that distinct latent information is present within unstructured text. Specifically, the distribution of scores generated by the Longformer model exhibited peaks at neutral and poor sentiment. Direct estimation of LOS by the LLM demonstrated similar peaks. Both distributions were significantly related to actual LOS and were able to capture approximately 15-20% of the variation in actual LOS based on our regression models.

In this context, Longformer has several advantages over other models. As an encoder-based model, it is designed to handle long document context. This allows it to string together key phrases throughout an admission note, likely improving detection of sentiment that would be difficult to detect in isolated fragments of the notes. Models with more limited scopes (VADER, TextBlob) are unable to synthesize these fragments. Meanwhile, LLMs such as the GPT-oss 20-B model can effectively compile information in longer admission notes, but require huge computational overhead to do so. While not accurate enough to serve as a standalone LOS predictor, our work suggests relatively computationally inexpensive models like Longformer could serve as viable adjuncts, capable of extracting hidden information regarding disease severity that might be missed by structured data alone.

### 4.3 LLMs and Zero-Shot Estimation

Notably, but perhaps unsurprisingly, prompting the GPT-oss 20B model for direct estimation of LOS outperformed the same model prompted as a sentiment predictor. This result demonstrates the additional element of variability in LLM approaches introduced by prompt language. By asking the model to act as a medical administrator predicting LOS, we forced the LLM to map the text to clinical severity rather than emotional tone. No matter how sophisticated the prompt, however, relying on zero-shot inference likely severely limits the model’s potential. Zero-shot prompting asks the model to perform a complex, domain-specific task without providing any task-specific examples or training weights updated for the target domain. Substantial performance gains are likely possible via supervised fine-tuning (e.g., Low-Rank Adaptation), access to outside information (e.g., Retrieval Augmented Generation), and chain-of-thought prompting. Such techniques have shown promise in related tasks such as prediction of operative risk and post-operative length of stay^14,15^.

### 4.4 Comparison with Existing Approaches

Although our results indicate information contained within unstructured text has a small but measurable association with LOS, the existing literature strongly suggests structured clinical data remains the superior modality for LOS estimation. Machine learning methods using structured data such as sex, age, BMI, vital signs, medications, and initial laboratory results have set a high benchmark, achieving concordance indices of over 0.85^16^. Still, most existing models focus on binary (e.g. short vs. long LOS) outcomes rather than continuous prediction of LOS, limiting their utility for hospital bed management.

## 5. Conclusion

In conclusion, while sentiment analysis reveals a statistically significant link to hospital duration, its practical utility is hampered by the objective nature of clinical language. In many contexts, LLMs have shown promise in solving clinically complex issues, but our results demonstrate limited success in zero-shot prediction of LOS from admission note text alone. Future research should focus on multimodal models that integrate the high-performing structured variables identified in the literature with fine-tuned models capable of extracting latent and otherwise unutilized information from unstructured clinical narratives.

## Data Availability

Aggregate data produced in the study are available upon reasonable request to the authors. Patient level data is not available due to privacy concerns.

## Notes

### Competing Interest Statement

The authors have declared no competing interest.

### Funding Statement

This study did not receive any funding.

### Author Declarations

The IRB of Baylor College of Medicine waived ethical approval for this work.

